# Clinical Efficacy of Intravenous Immunoglobulin Therapy in Critical Patients with COVID-19: A Multicenter Retrospective Cohort Study

**DOI:** 10.1101/2020.04.11.20061739

**Authors:** Ziyun Shao, Yongwen Feng, Li Zhong, Qifeng Xie, Ming Lei, Zheying Liu, Conglin Wang, Jingjing Ji, Liu Huiheng, Zhengtao Gu, Zhongwei Hu, Lei Su, Ming Wu, Zhifeng Liu

**Author notes:** To whom correspondence should be addressed: Dr. Zhifeng Liu, Department of Critical Care Medicine, General Hospital of Southern Theatre Command of PLA, Guangzhou, 510010, China., Dr. Ming Wu, Department of Critical Care Medicine and Hospital Infection Prevention and Control, The Second People’s Hospital of Shenzhen & First Affiliated Hospital of Shenzhen University, Health Science Center, Shenzhen, 518035, China., Prof. Lei Su, Department of Critical Care Medicine, General Hospital of Southern Theatre Command of PLA, Guangzhou, 510010., Prof. Zhongwei Hu, Department of Nephrology, Guangzhou Eighth people’s hospital, Guangzhou Medical University, Guangzhou, 510060, China. These authors contributed equally to this work (Ziyun Shao, Yongwen Feng, Li Zhong, Qifeng Xie, Ming Lei).

## Abstract

**Importance:** Coronavirus disease 2019 (COVID-19) has become pandemic, causing more than 1.5 million infections and over ten-thousands of deaths in a short period of time worldwide. However, little is known about its pathological mechanism, and reports on clinical study on specific treatment are few.

**Objective:** The purpose of this study is to determine the clinical efficacy of intravenous immunoglobulin (IVIG) therapy in COVID-19 patients.

**Design, setting and participants:** This multicenter retrospective cohort study enrolled 325 adult critical COVID-19 patients, including severe type and critical type, according to the clinical classification defined by National Health Commission of China, in 8 government designated treatment centers in China from Dec 23, 2019 to Mar 31, 2020. Demographic, clinical, treatment, and laboratory data as well as prognosis were extracted from electronic medical records.

**Exposure:** IVIG was exposure factor.

**Main outcomes and measures:** Primary outcomes were the 28-day and 60-day mortality, and secondary outcomes were the total length of in-hospital and the total duration of the disease. Meanwhile, the parameters of inflammation responses and organ functions were measured. The risk factors were determined by COX proportional hazards model. The subgroup analysis was carried out according to clinical classification of COVID-19, IVIG dosage, and timing.

**Results:** In the enrolled 325 patients, 222 (68%) were severe type and 103 (32%) were critical type; 42 (13%) died in 28-day within hospitalization, and 54 (17%) died within 60-day; The death in 60-day includes 6 (3%) severe type patients and 48 (47%) critical type patients. 174 cases were used IVIG, and 151 cases were not. Compared with the baseline characteristics between two groups, the results showed that the patients in IVIG group presented higher Acute Physiology and Chronic Health Evaluation (APACHII) score and Sequential Organ Failure Assessment (SOFA) score, higher plasm levels of IL-6 and lactate, and lower lymphocyte count and oxygenation index (all *P*<0.05). The 28-day and 60-day mortality were not improved with IVIG in overall cohort. The in-hospital stay and the total duration of disease were longer in IVIG group (*P*<0.001). Risk factors were clinical classifications (hazards ratio 0.126, 95% confidence interval 0.039-0.413, *P*=0.001), and using IVIG (hazards ratio 0.252, 95% confidence interval 0.107-0.591, *P*=0.002) with COX proportional hazards model. Subgroup analysis showed that only in patients with critical type, IVIG could significantly reduce the 28-day mortality, decrease the inflammatory response, and improve some organ functions (all *P*<0.05); and application of IVIG in the early stage (admission≤7 days) with a high dose (>15 g/d) exhibited significant reduction of 60-day mortality in the critical type patients.

**Conclusions and Relevance:** Early administration of IVIG with high dose improves the prognosis of critical type patients with COVID-19. This study provides important information on clinical application of the IVIG in treatment of SARS-CoV-2 infection, including patient selection and administration timing and dosage.

**Key points:** *Question:* Intravenous immunoglobulin (IVIG) was recommended to treat critical Coronavirus disease 2019 (COVID-19) patients in a few reviews, but the clinical study evidence on its efficacy in COVID-19 patients was lacked.

*Finding:* In this multicenter cohort study that included 325 adult critical patients from 8 treatment centers, the results showed that early administration (admission ≤ 7 days) of IVIG with high dose (> 15 g/d) improves the prognosis of critical type patients with COVID-19.

*Meaning:* This study provides important information on clinical application of IVIG in treatment of SARS-CoV-2 infection, including patient selection, administration timing and dosage.

## Introduction

Coronavirus disease-2019(COVID-19) is a systemic infectious disease mainly caused by severe acute respiratory syndrome coronavirus 2 (SARS-CoV-2), while critical COVID-19 is a life-threatening multi-organ dysfunction syndrome dysregulated resulted from host response to SARS-CoV-2 and characterized by refractory hypoxemia caused by acute respiratory distress syndrome (ARDS) [1]. From December 2019 to April 2020, more than 80,000 people in China infected SARS-CoV-2, and in which, over 3,000 people died. Globally, more than 1.5 million people infected SARS-CoV-2 and near 100,000 people died, including a large number of health workers, which has become the most serious problem faced by all medical staff and researchers [2, 3]. According to the reports, the general mortality was about 1%-15% in all COVID-19 cases, and the incidence of critical COVID-19, including both severe and critical types defined by Chinese Recommendations for Diagnosis and Treatment of Novel Coronavirus (SARS-CoV2) Infection published by National Health Commission of China (Trial 7th Version), is about 10%− 20%, and its mortality was about 30%-60% [4, 5].

It is currently believed that SARS-CoV-2 primarily infect the lungs, and subsequently cause systemic inflammation and immune response disorder, and ultimately lead to multiple organ injury and even death [6]. However, effective therapeutic method is lacking.

The available clinical treatment strategies to critical COVID-19 are mainly antiviral and oxygen therapy, as well as organ and symptomatic support, including mechanical ventilation, even extracorporeal membrane oxygenation (ECMO) of cardiopulmonary support, and continuous renal replacement therapies (CRRT) [7]. However, the clinical efficacy of these strategies is still uncertain. Some clinical tests and autopsy results suggested that the inflammation and immune response caused by the virus infection is the key factor to the progress of disease and the poor prognosis, but the underlying mechanisms remain unclear [8,9]. Targeted intravenous immunoglobulin (IVIG) is one of the main treatment measures [4, 7]. However, due to the lack of clinical trials, the efficacy of IVIG has yet to be determined and clinical application is still controversial.

Human Immunoglobulin (pH4) for intravenous injection is a liquid preparation containing human immunoglobulins made from normal human plasma, containing IgG antibody with broad-spectrum antiviral, bacterial or other pathogens. IVIG can rapidly increase the IgG level in the blood, directly neutralize exogenous antigens, and regulate multiple immune functions, including regulating immune media, and improving the immune capacity of natural immune cells and lymphocytes. IVIG has been wildly used in the treatment of severe bacterial and viral infection and sepsis [10, 11]. Some studies demonstrated its clinical efficacy, especially in the case of viral infectious diseases [12]; whereas, other studies failed to show its therapeutic efficacy, leading to great controversy on its clinical application in acute respiratory virus infection [13]. The latest version of China therapeutic guidelines of COVID-19 suggested IVIG as a selective treatment method. However, due to lacking specific antibodies against SARS-CoV-2, the efficacy of IVIG remains to be elucidated. It is noticed that use of IVIG is recommendation in the list of selective methods in the COVID-19 therapeutic guidelines of WHO [4,7,14,15].

In order to confirm the potential therapeutic efficacy of IVIG to COVID-19, we retrospective collected the clinical and outcome data of critical COVID-19 patients, including both severe type and critical type, from 8 government designated treatment centers in three cities (Wuhan, Guangzhou, and Shenzhen) in China, and using IVIG as an exposure factor analyzed the symptoms and outcomes. Up to date, this is the first clinical multicenter cohort study on IVIG treatment for COVID-19 with a large number of critical ill patients. This study provides important information on clinical application of the IVIG in treatment of SARS-CoV-2 infection, including patient selection and administration timing and dosage.

## Methods

### Study design and participants

This multicenter retrospective cohort study was performed in eight government designated treatment centers for COVID-19 patients (4 ICUs and 4 general wards) in 3 cities in China, including Wuhan, Guangzhou, and Shenzhen. The data collection period was from December 2019 to March 2020, and the data cutoff date was April 3, 2020.

Inclusion Criteria: (1) Adult aged >=18 years old; (2) Laboratory (RT-PCR) confirmed SARS-COV-2 infection in throat swab and/or sputum and/or lower respiratory tract samples; or conformed plasma positive of specific antibody (IgM or/and IgG) against SARS-COV-2; (3)

In-hospital treatment ≥72 hours; (4) Meet any one of the following a-c criteria for severe type or d-f criteria for critical type: (a) Respiratory rate >=30/min; or (b) Rest SPO2<=90%; or (c) PaO2/FiO2<=300 mmHg; or (d) Respiratory failure and needs mechanical ventilation; or (e) Shock occurs; or (f) Multiple organ failure and needs ICU monitoring;

Exclusion Criteria: (1) Exist of other evidences that can explain pneumonia including but not limited to influenza A virus, influenza B virus, bacterial pneumonia, fungal pneumonia, noninfectious causes, etc.; (2) Women who are pregnant or breast-feeding; (3) Researchers consider unsuitable.

### Procedures

We designed the data collection form, which includes the demographic, clinical, treatment, laboratory data and prognosis were extracted from electronic medical records. Detailed clinical data before and after prescription IVIG, and the data at the corresponding time of the same period in non-IVIG group were collected, respectively. Whether and when to use IVIG, dosage and course were decided by the doctors in charge. Comparison was conducted according to whether IVIG was used or not. Primary endpoint was 28-day and 60-day in hospital mortality, and total in-hospital days and the total duration of the disease as the secondary endpoint. Meanwhile, the parameters of inflammation and organ function were measured. The influencing factors were determined by COX regression. Analysis of the outcome and the survival curves were carried out according to clinical classification of COVID-19, IVIG dosage, and timing. The study was approved by the Research Ethics Commission of General Hospital of Southern Theater Command of PLA (HE-2020-08) and the requirement for informed consent was waived by the Ethics Commission.

### Definitions

“Critical COVID-19” in this article is defined as combined term of “Severe type” and “Critical type” of COVID-19, classified following Chinese Recommendations for Diagnosis and Treatment of Novel Coronavirus (SARSCoV2) infection (Trial 7th version) published by National Health Commission of China. IVIG represents the human Immunoglobulin for intravenous injection, which is a liquid preparation containing human immunoglobulins made from normal human plasma, containing IgG antibody with broad-spectrum anti-viral, bacterial, or other pathogens. IVIG rapidly increases the level of IgG in the blood of the recipient after intravenous infusion, and enhance the anti-infection ability and immune regulation function of the body.

### Statistical Analysis

The categorical data were summarized as numbers and percentages, and inter-group comparisons were performed using Mann-Whitney U, χ2 tests or Fisher’s exact test. Continuous variables were expressed as the arithmetic mean and standard deviation (SD) or as the median and interquartile range, depending on whether or not they showed a gaussian distribution. Continuous data with gaussian distribution were compared with the Student’s t test or one-way ANOVA and those with a non-gaussian distribution, with the Wilcoxon rank-sum test. To compare the white blood cell, lymphocytes, neutrophils, and monocyte count between IGIV and non-IGIV groups. To determine the independent effect of 60-day mortality in critical COVID-19 patients after accounting for significant confounders, COX proportional hazards model was used with fully adjusted model: OR (odds ratio) and 95% confidence interval levels (95% CI).

Moreover, for analysis of the 28-day and 60-day mortality, Kaplan-Meier Survival Curves and the Log-Rank Test were used. Statistical analysis was performed using the SPSS Windows version 11.0 statistical package (SPSS Inc, Chicago, IL), *P* values (two-tailed) below 0.05 were considered statistically significant.

### Role of the funding source

The funder of the study had no role in study design, data collection, data analysis, data interpretation, or writing of the report. The corresponding author had full access to all the data in the study and had final responsibility for the decision to submit for publication.

## Results

### Demographics and baseline characteristics

Clinical data of 338 patients were collected with confirmed critical COVID-19. After excluding 13 patients due to missing key information, 325 patients were included in the final analysis (Figure 1). The detailed demographic and clinical profile data of all critical ill patients with COVID-19 on baseline were summarized in Table 1. Patient’s mean age was 58 ys (IQR 46.0-69.0), mean body temperature was 37.0°C (IQR 36.5-37.8). Nearly half of the patients had comorbidity, mainly hypertension (30%), diabetes (12%) and coronary heart disease (10%). 222 (68%) were severe type and 103 (32%) were critical type. 174 cases used IVIG, and 151 cases were not. Comparisons of baseline characteristics between two groups showed that the disease was more severe in the IVIG group, presented by older age, higher APACHII scores and SOFA scores, higher levels of total bilirubin, direct bilirubin, creatinine, c-reactive protein, IL-6, and lactate, but lower platelets and lymphocyte count (all *P*<0.05), and decreased PaO_2_/FiO_2_ (*P*=0.011, Table 1).

**Table 1:**
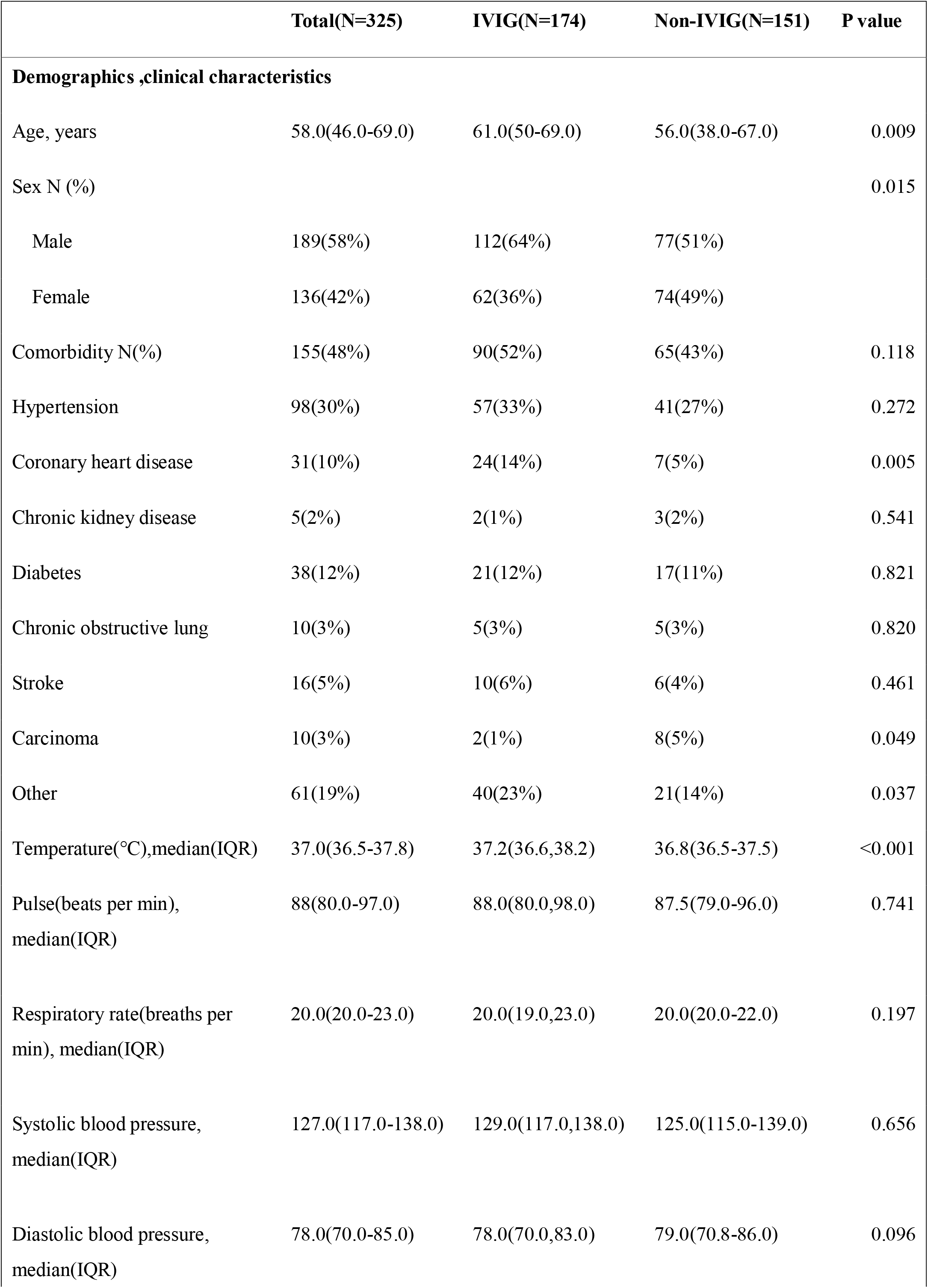
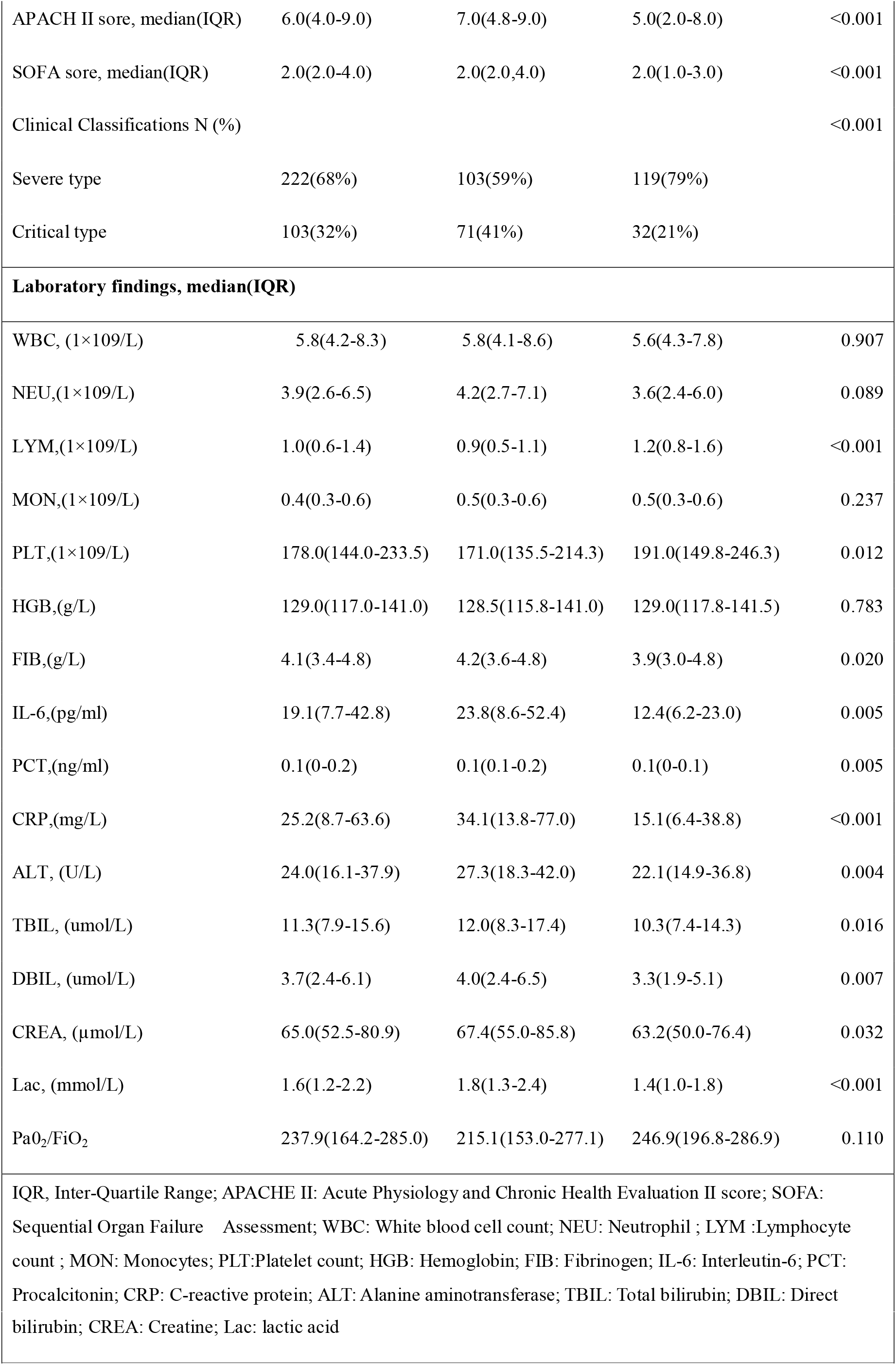
Baseline characteristics of demographics, clinical and laboratory findings in IVIG group and Non-IVIG group.

|  | **Total(N=325)** | **IVIG(N=174)** | **Non-IVIG(N=151)** | **P value** |
| --- | --- | --- | --- | --- |
| **Demographics ,clinical characteristics** |  |  |  |  |
| Age, years | 58.0(46.0-69.0) | 61.0(50-69.0) | 56.0(38.0-67.0) | 0.009 |
| Sex N (%) |  |  |  | 0.015 |
| Male | 189(58%) | 112(64%) | 77(51%) |  |
| Female | 136(42%) | 62(36%) | 74(49%) |  |
| Comorbidity N(%) | 155(48%) | 90(52%) | 65(43%) | 0.118 |
| Hypertension | 98(30%) | 57(33%) | 41(27%) | 0.272 |
| Coronary heart disease | 31(10%) | 24(14%) | 7(5%) | 0.005 |
| Chronic kidney disease | 5(2%) | 2(1%) | 3(2%) | 0.541 |
| Diabetes | 38(12%) | 21(12%) | 17(11%) | 0.821 |
| Chronic obstructive lung | 10(3%) | 5(3%) | 5(3%) | 0.820 |
| Stroke | 16(5%) | 10(6%) | 6(4%) | 0.461 |
| Carcinoma | 10(3%) | 2(1%) | 8(5%) | 0.049 |
| Other | 61(19%) | 40(23%) | 21(14%) | 0.037 |
| Temperature(°C),median(IQR) | 37.0(36.5-37.8) | 37.2(36.6,38.2) | 36.8(36.5-37.5) | <0.001 |
| Pulse(beats per min),<br>median(IQR) | 88(80.0-97.0) | 88.0(80.0,98.0) | 87.5(79.0-96.0) | 0.741 |
| Respiratory rate(breaths per<br>min), median(IQR) | 20.0(20.0-23.0) | 20.0(19.0,23.0) | 20.0(20.0-22.0) | 0.197 |
| Systolic blood pressure,<br>median(IQR) | 127.0(117.0-138.0) | 129.0(117.0,138.0) | 125.0(115.0-139.0) | 0.656 |
| Diastolic blood pressure,<br>median(IQR) | 78.0(70.0-85.0) | 78.0(70.0,83.0) | 79.0(70.8-86.0) | 0.096 |
| APACH II sore, median(IQR) | 6.0(4.0-9.0) | 7.0(4.8-9.0) | 5.0(2.0-8.0) | <0.001 |
| SOFA sore, median(IQR) | 2.0(2.0-4.0) | 2.0(2.0,4.0) | 2.0(1.0-3.0) | <0.001 |
| Clinical Classifications N (%) |  |  |  | <0.001 |
| Severe type | 222(68%) | 103(59%) | 119(79%) |  |
| Critical type | 103(32%) | 71(41%) | 32(21%) |  |
| **Laboratory findings, median(IQR)** |  |  |  |  |
| WBC, (1×10<sup>9</sup>/L) | 5.8(4.2-8.3) | 5.8(4.1-8.6) | 5.6(4.3-7.8) | 0.907 |
| NEU,(1×10<sup>9</sup>/L) | 3.9(2.6-6.5) | 4.2(2.7-7.1) | 3.6(2.4-6.0) | 0.089 |
| LYM,(1×10<sup>9</sup>/L) | 1.0(0.6-1.4) | 0.9(0.5-1.1) | 1.2(0.8-1.6) | <0.001 |
| MON,(1×10<sup>9</sup>/L) | 0.4(0.3-0.6) | 0.5(0.3-0.6) | 0.5(0.3-0.6) | 0.237 |
| PLT,(1×10<sup>9</sup>/L) | 178.0(144.0-233.5) | 171.0(135.5-214.3) | 191.0(149.8-246.3) | 0.012 |
| HGB,(g/L) | 129.0(117.0-141.0) | 128.5(115.8-141.0) | 129.0(117.8-141.5) | 0.783 |
| FIB,(g/L) | 4.1(3.4-4.8) | 4.2(3.6-4.8) | 3.9(3.0-4.8) | 0.020 |
| IL-6,(pg/ml) | 19.1(7.7-42.8) | 23.8(8.6-52.4) | 12.4(6.2-23.0) | 0.005 |
| PCT,(ng/ml) | 0.1(0-0.2) | 0.1(0.1-0.2) | 0.1(0-0.1) | 0.005 |
| CRP,(mg/L) | 25.2(8.7-63.6) | 34.1(13.8-77.0) | 15.1(6.4-38.8) | <0.001 |
| ALT, (U/L) | 24.0(16.1-37.9) | 27.3(18.3-42.0) | 22.1(14.9-36.8) | 0.004 |
| TBIL, (umol/L) | 11.3(7.9-15.6) | 12.0(8.3-17.4) | 10.3(7.4-14.3) | 0.016 |
| DBIL, (umol/L) | 3.7(2.4-6.1) | 4.0(2.4-6.5) | 3.3(1.9-5.1) | 0.007 |
| CREA, (μmol/L) | 65.0(52.5-80.9) | 67.4(55.0-85.8) | 63.2(50.0-76.4) | 0.032 |
| Lac, (mmol/L) | 1.6(1.2-2.2) | 1.8(1.3-2.4) | 1.4(1.0-1.8) | <0.001 |
| PaO<sub>2</sub>/FiO<sub>2</sub> | 237.9(164.2-285.0) | 215.1(153.0-277.1) | 246.9(196.8-286.9) | 0.110 |
| IQR, Inter-Quartile Range; APACHE II: Acute Physiology and Chronic Health Evaluation II score; SOFA: Sequential Organ Failure Assessment; WBC: White blood cell count; NEU: Neutrophil ; LYM :Lymphocyte count ; MON: Monocytes; PLT:Platelet count; HGB: Hemoglobin; FIB: Fibrinogen; IL-6: Interleutin-6; PCT: Procalcitonin; CRP: C-reactive protein; ALT: Alanine aminotransferase; TBIL: Total bilirubin; DBIL: Direct |  |  |  |  |

|  |
| --- |
| bilirubin; CREA: Creatine; Lac: lactic acid |
| **Table 1: Baseline characteristics of demographics, clinical and laboratory findings in**<br>**IVIG group and Non-IVIG group** |

**Figure 1:**
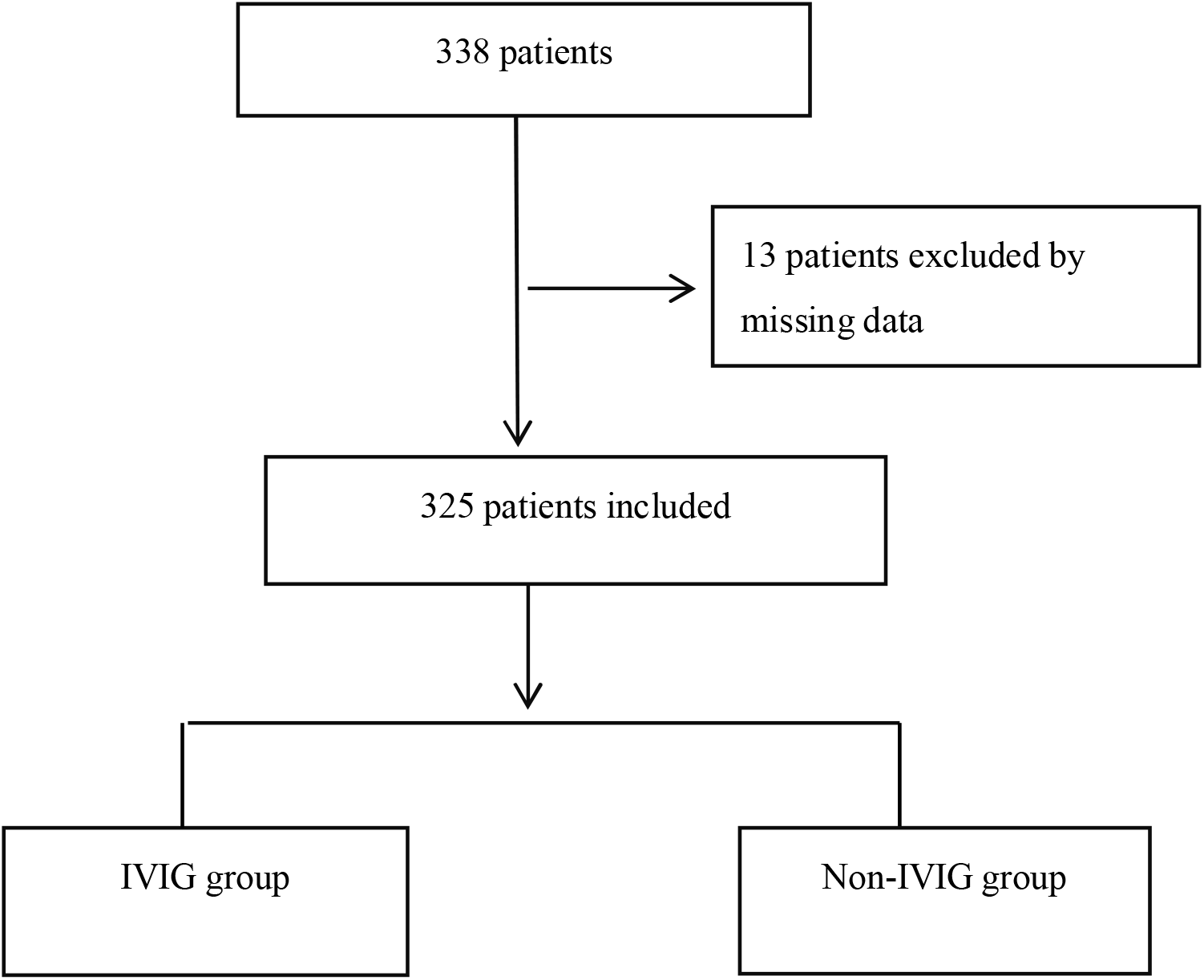
Flow chart of all excluded and included patients.

### Primary and secondary outcomes in all patients

Analysis of primary and secondary outcomes in all patients showed that 42 (13%) died in 28 days and 54 (17%) died in 60 days; Death in 60-day includes 6 (3%) severe type patients and 48 (47%) critical type patients. In IVIG group 22 (13%) died within 28-day, total 33 (19%) died in 60-day. In non-IVIG group 20 (13%) died within 28-day, total 21 (14%) died in 60-day. There was no significant difference in 28-day and 60-day mortality between the IVIG group and non-IVIG group (*P*=0.872 and *P*=0.222, respectively), and no significant difference in survival time (*P*= 0.225, table 2, supplementary Figure1). Analysis of secondary outcomes in all patients showed that the median time of in hospital stay was 20.0 days (IQR 14.0-28.0), and the total course of disease was 28.0 days (19.0-37.0). Compared between the two groups, the in hospital day and total duration of disease were longer in IVIG group (both *P* < 0.001, table 2), and the number of lymphocytes was still lower (*P* < 0.001), CRP was still higher (*P* = 0.011, supplementary table 1), which is consistent with the more serious initial condition of IVIG group.

**Table 2:** Primary and secondary outcomes in IVIG and Non IVIG groups.

|  | **Total(N=325)** | **IVIG(N=174)** | **Non-IVIG(N=151)** | **P value** |
| --- | --- | --- | --- | --- |
| **Primary outcomes N(%)** |  |  |  |  |
| 28-day mortality | 42(13%) | 22(13%) | 20(13%) | 0.872 |
| 60-day mortality | 54(17%) | 33(19%) | 21(14%) | 0.222 |
| **Secondary outcome , median(IQR)** |  |  |  |  |
| In-hospital days | 20.0(14.0-28.0) | 23.5(16.0-33.0) | 16.0(13.0-22.0) | <0.001 |
| Total course of disease <sup>a</sup> | 28.0(19.0-37.0) | 31.0(23.0-39.0) | 23.0(17.0-31.0) | <0.001 |
| <sup>a</sup>Total course of disease : Time from illness onset to death or discharge, days |  |  |  |  |
| **Table 2: Primary and secondary outcomes in IVIG and Non IVIG groups** |  |  |  |  |

### Risk factors analysis for 60 day in-hospital mortality

In order to correct the bias of the difference between the two groups’ basic conditions on the prognosis, multivariable COX regression analysis was performed with gender, age, comorbidity, APACHE II score, SOFA score, temperature, white blood cell count, neutrophil count, lymphocyte count, fibrinogen, creatinine, PaO_2_/FiO_2_, lactic acid, clinical classification of COVID-19, and IVIG use. It was found that lower lymphocyte count associated with more severe COVID-19 classification and a higher 60-day mortality, while, application of IVIG significantly decreased the 60-day mortality (HR 0.252; 95% CI 0.107-0.591; *P*=0.002, Table 3).

**Table 3:** Multivarate analysis for factors associated with death in hospital.

| Variable | Univariable | P value | Multivariate | P value |
| --- | --- | --- | --- | --- |
|  | HR ( 95%CI ) |  | HR ( 95%CI ) |  |
| Gender | 1.253 ( 0.721-2.177 ) | 0.424 | 0.867(0.414-1.815) | 0.704 |
| Age | 1.057 ( 1.038-1.077 ) | <0.001 | 1.015(0.981-1.051) | 0.394 |
| Comorbidity | 2.562 ( 1.442-4.550 ) | 0.001 | 4.281(1.492-12.282) | 0.007 |
| APACHEII score | 1.143 ( 1.117-1.169 ) | <0.001 | 1.051(0.965-1.145) | 0.256 |
| SOFA score | 1.186 ( 1.151-1.221 ) | <0.001 | 1.125(0.988-1.281) | 0.076 |
| Temperature,°C | 0.762 ( 0.537-1.082 ) | 0.129 | N/A |  |
| White blood cell count | 1.157 ( 1.116-1.200 ) | <0.001 | N/A |  |
| Neutrophil | 1.167 ( 1.126-1.209 ) | <0.001 | N/A |  |
| Lymphocyte | 0.160 ( 0.074-0.347 ) | <0.001 | 0.315(0.116-0.859) | 0.024 |
| Fibrinogen | 0.955 ( 0.742-1.228 ) | 0.720 | N/A |  |
| Creatine | 1.009 ( 1.006-1.012 ) | <0.001 | N/A |  |
| PaO<sub>2</sub>/FiO<sub>2</sub> | 0.992 ( 0.988-0.996 ) | <0.001 | 0.998(0.994-1.002) | 0.395 |
| lactic acid | 1.022 ( 0.839-1.244 ) | 0.832 | 0.893(0.734-1.078) | 0.260 |
| Clinical classification | 0.045 ( 0.019-0.105 ) | <0.001 | 0.126(0.039-0.413) | 0.001 |
| IVIG | 1.371 ( 0.793-2.370 ) | 0.258 | 0.252(0.107-0.591) | 0.002 |
| APACHE II : Acute Physiology and Chronic Health Evaluation II score; SOFA: Sequential Organ Failure Assessment; |  |  |  |  |
| **Table 3: Multivariate analysis for factors associated with death in hospital** |  |  |  |  |

### Primary and secondary outcomes in subgroups

According to the results of multivariate analysis, deep analysis was carried out in different subgroups. The results showed that IVIG could significantly decrease the 28-day mortality of patients in critical type (*P*=0.009) but had no effects on the 60-day mortality and in-hospital stay (both *P*>0.05, Table 4). In addition, IVIG also significantly decreased the procalcitonin (PCT) and lactic acid levels in critical type patients (*P* < 0.05, supplementary Table 2), suggesting that IVIG decreases the inflammatory response and may improve microcirculation perfusion. However, in the severe type patients, there was no difference in mortality between IVIG group and non IVIG group (*P*> 0.05), and the length of in-hospital stay in IVIG group was not changed (Table 4). Moreover, there was no difference in the 60-day survival rate between the two groups (supplementary Figure 2 and 3).

**Table 4:**
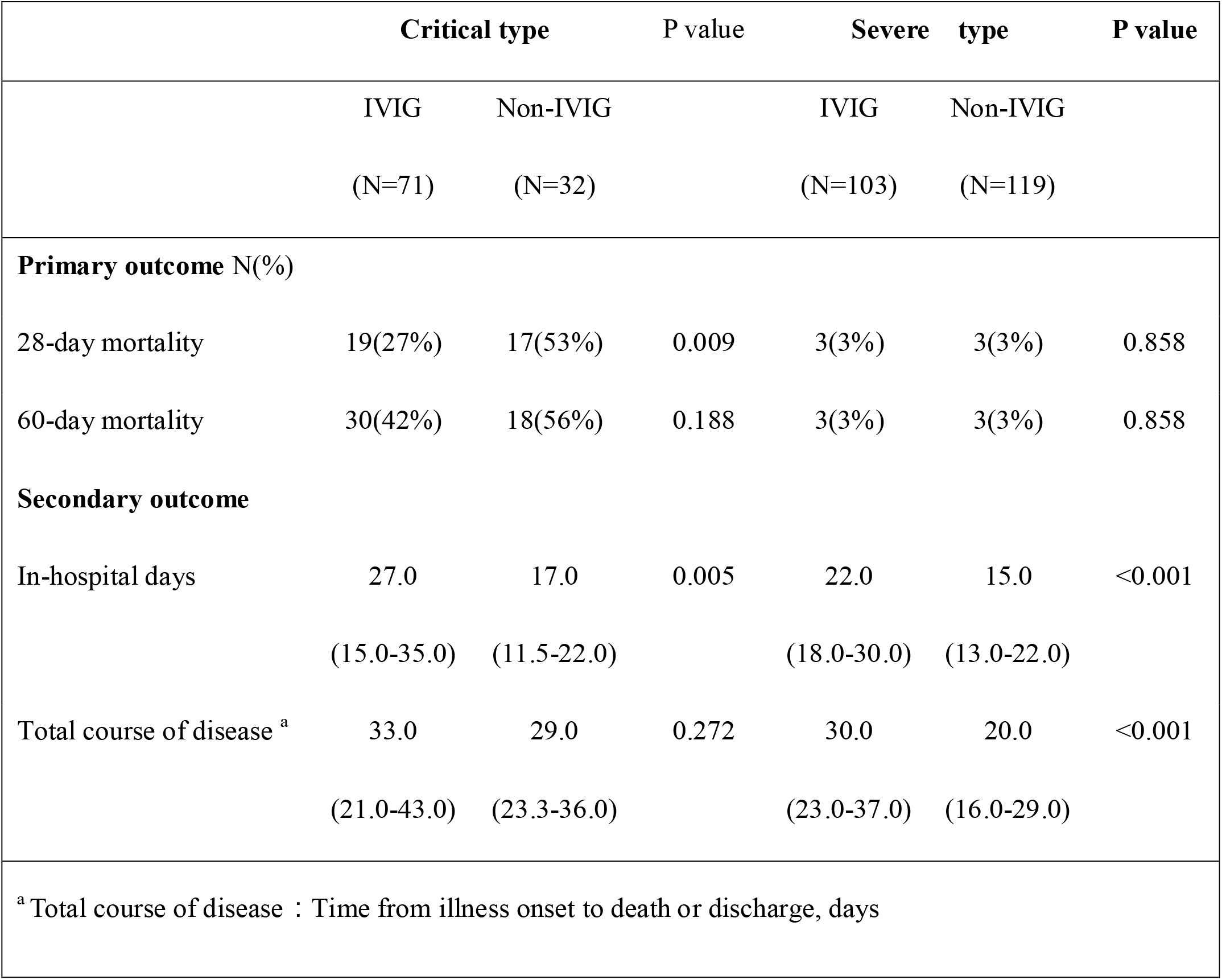
Effects of IVIG treatment on primary and secondary outcome analysis in subgroup of critical and severe type.

Further analyses were performed with APACHII score (> 11 and ≦11), PaO_2_ / FiO_2_ (> 100 and ≦100), lymphocyte count (≦0.8×10^9^/L and >0.8×10^9^/L), and SOFA score (≦6 and > 6), and the results showed that there were no differences in 28-day, 60-day mortality between IVIG and non IVIG groups (supplementary Table 3). In the comparison of parameters of inflammation response and organ function, IVIG significantly decreases PCT level and improves PaO_2_/FiO_2_ (both *P*< 0.05, supplementary Table 4) in patients with APACHII score >11 or lymphocyte count ≤0.8×10^9^/L.

To further confirm the effects of IVIG dosage on the outcomes of COVID-19 patients, subgroup with different doses of IVIG (>15 g/d and ≤15 g/d) were compared, and the results showed that high dose IVIG (>15 g/d) significantly reduces 28-day and 60-day mortality (*P*= 0.044, 0.049, respectively, Table 5), and increases survival time (*P*= 0.045, supplementary Figure 4) as compared with the low dose group (≤15 g/d). Further comparison between the subgroups with different COVID-19 types showed that high dose IVIG of more than 15 g/d could significantly reduce the 28-day and 60-day mortality of the critical type patients (*P* = 0.002, *P* < 0.0001, perspective, Table 6), and increase the survival time (*P* < 0.0001, supplementary Figure 5). In contrast, in the severe type patients, neither high nor low dose of IVIG demonstrated effects (Table 6, supplementary Figure 6). By comparing the effects of different doses on the parameters of inflammatory response and organ functions, the results showed that high doses of IVIG (>15g/d) could significantly decrease the lactic level and increase PaO_2_/FiO_2_ (both *P* < 0.05, supplementary Table 5), especially in the critical type patients (supplementary Table 6).

**Table 5:** Effects of different dose of IVIG treatment on primary and secondary outcomes in all patients.

|  | Total<br>(N=174) | IVIG>15g/d<br>(N=74) | IVIG≤15g/d<br>(N=100) | P value |
| --- | --- | --- | --- | --- |
| **Primary outcomes** N(%) |  |  |  |  |
| 28-day mortality | 22(13%) | 5(7%) | 17(17%) | 0.044 |
| 60-day mortality | 33(19%) | 9(12%) | 24(24%) | 0.049 |
| **Secondary outcome** , median(IQR) |  |  |  |  |
| In-hospital days | 23.5(16.0-33.0) | 26.5(18.0-33.0) | 22.0(16.0-31.0) | 0.091 |
| Total course of disease | 31.0(23.0-39.0) | 32.0(24.0-39.0) | 30.0(22.0-39.0) | 0.517 |

**Table 6:**
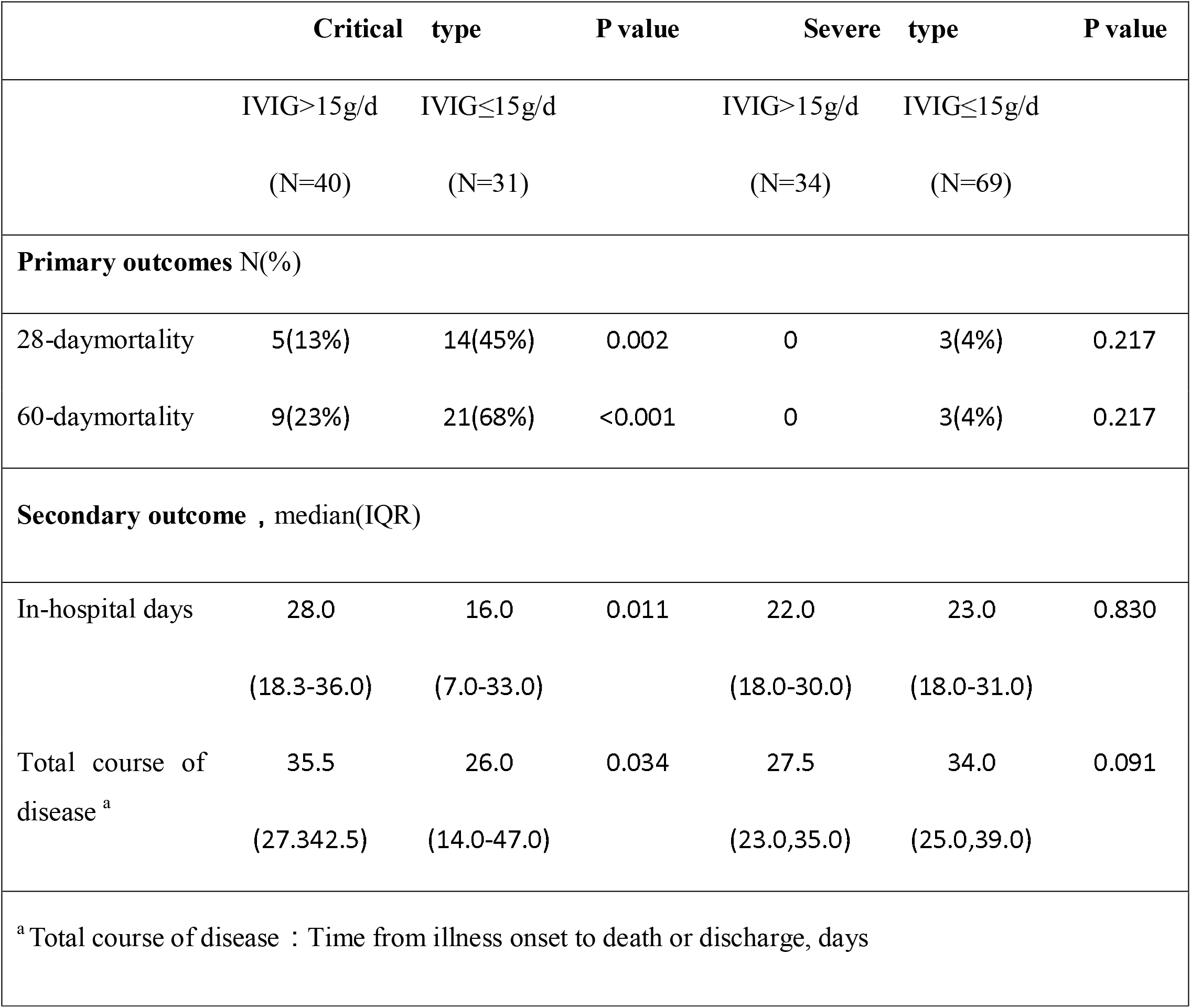
Effects of different dose of IVIG on primary and secondary outcome in critical and severe subgroup.

To further confirm the effects of IVIG application timing on the outcomes of COVID-19 patients, subgroup with the time from admission to the beginning of IVIG treatment (>7 d and ≤ 7 d admission) were compared, and the results showed that early administration of IVIG (≤ 7d) could significantly reduce 60-day mortality (*P*= 0.008, Table 7), total in-hospital stay, and total course of disease (*P*=0.025 and *P*=0.005, respectively), and significantly increase survival time (supplementary Figure 7). Further analysis showed that early use of IVIG could also significantly increase PaO_2_/FiO_2_ (*P*= 0.022) and decrease levels of PCT (*P*=0.016), IL-6, and CRP (supplementary Table 7), suggesting that early use of IVIG is potentially able to improve inflammatory response and some organ functions.

**Table 7:** Effects of the timing of IVIG use on the primary and secondary outcome.

|  | Total<br>(N=174) | IVIG>7d<br>(N=16) | IVIG≤7d<br>(N=158) | P value |
| --- | --- | --- | --- | --- |
| **Primary outcomes** N(%) |  |  |  |  |
| 28-day mortality | 22(13%) | 3(19%) | 19(12%) | 0.441 |
| 60-day mortality | 33(19%) | 7(44%) | 26(17%) | 0.008 |
| **Secondary outcome** , median(IQR) |  |  |  |  |
| In-hospital days | 23.5(16.0-33.0) | 31.0(23.0-39.8) | 22.0(16.0-32.0) | 0.025 |
| Total course of disease <sup>a</sup> | 31.0(23.0-39.0) | 41.5(31.0-49.0) | 30.0(23.0-38.0) | 0.005 |
| <sup>a</sup> Total course of disease : Time from illness onset to death or discharge, days |  |  |  |  |

## Discussion

The pandemic outbreak of COVID-19 is rapidly spreading all over the world, resulting in over one hundred thousand global death due to lacking well-established treatment. To our knowledge, this multicenter retrospective cohort study is the first clinical evaluation, with a large number of cases, on the efficiency of IVIG treatment to critical COVID-19 patients. The basic condition of patients in IVIG group was more serious. The results showed that, for the critical COVID-19 patients, IVIG has no effect on the 28-day and 60-day mortality. Notably, multivariable regression showed that both classification of COVID-19 and IVIG using were the factors that related with hazards ratios of death. Subgroup analysis showed that only in the critical type patients IVIG could significantly decrease the inflammatory response, improve some organ functions, reduce the 28-day mortality rate, and prolong the survival time. Furthermore, the study showed that early use of IVIG (admission≤ 7 days) with high dose (>15 g/d) exhibits more significantly curative effect. Noteworthy, the results indicated that early and high dose of IVIG therapy seems only effective in the critical type patients showing an improved prognosis. These findings provide important information on clinical application of the IVIG in treatment of SARS-CoV-2 infection, including patient selection and administration timing and dosage.

Regarding the clinical application of immunoglobulin in COVID-19 patients, including the efficacy and way to use (timing and dosage), is still controversial. Pharmacological studies suggested that high dose of IVIG pulse therapy leads to the formation of immunocomplex with pathogen antigen, which can be further cleared in circulation [12]. Immunoglobulin has been used in the treatment of virus infectious diseases, such as viral pneumonia, and autoimmune diseases [13]. In patients with severe COVID-19, but not in patients with mild disease, lymphopenia is a common feature, with drastically reduced numbers of CD4^+^ T cells, CD8^+^ T cells, B cells and natural killer (NK) cells [6, 24,25]. Immunoglobulin showed antiviral and anti-inflammatory effects through increasing certain cytokine secretion, such as IL-2, to promote T cell and B cell proliferation and differentiation, and therefore, immunoglobulin is thought to be beneficial in the treatment of COVID-19. Previous studies in the treatment of SARS and MERS suggested beneficial role for high dose immunoglobulin administration [16,17]. Since immunoglobulin was not the specific antibody to any virus and the clinical evidence for its efficacy was limited, some researchers held the oppose view about usage of immunoglobulin in acute virus infection [18,19]. In COVID-19 treatment guidance of China and WHO, the recommendation on immunoglobulin usage is different. The data from the current study showed that IVIG did not improve the outcome in overall enrolled critical COVID-19 patients. However, the multivariable regression and subgroup analysis showed that IVIG could only improve the prognosis in the critical type patients. One explanation was that severe type cases with lower organ injury and mortality, which is similar with previous study.

The recommended dose of immunoglobulin is 0.5 g/kg·d for 5 days. However, in the present study, the doses used differ among the different centers and physicians, ranging from 0.1 to 0.5 g/kg·d. The treatment period ranged from 5 to 15 days. By subgroup analysis, we found that only high dose over 15 g/d (equivalent to 0.2-0.3g/kg.d) shows the curative effect, which is consistent with the usage of immunoglobulin in treating sepsis only effectively in high dose[20]. High dose of immunoglobulin would be able to rapidly increase its concentration in circulation by 3- to 6-fold and play a role in enhancing immunity and antiviral function.

The current study suggests the importance of early use of immunoglobulin in the COVID-19 patients. Immunoglobulin affects both innate and adapted immune systems, and directly binds to pathogen antigen, which usually appears in the circulation in an early stage following virus infection [21,22, 23]. Based on the current understanding of the COVID-19 pathogenesis, in late stage, excessive inflammatory response is developed, and the organ dysfunction occurs, the efficacy of administration of immunoglobulin would be largely limited [8,9]. Our data showed that immunoglobulin employed within 7 days after hospital admission could improve the prognosis. And we found that patients in enrolled with IVIG group were in more severe condition, evidenced by higher APACHE II and SOFA scores, higher levels of IL-6 and lactate, as well as decreased lymphocyte count and oxygenation index in this multicenter retrospective study.

Compared to SARS and MERS, COVID-19 demonstrates several exceptionalities, including prolonged course, potential asymptomatic hypoxia, severe lung injury, and unexpected progress induced death [4,5,24]. These clinical features urgently call for exploratory treatment attempts. IVIG is one of the such attempts. To exclude the influence of the bias on the study, we performed the regression analysis on the potential factors. Univariate survival analysis showed that APACHE II and SOFA sores were the risk factor which related with the outcome. In further analysis, we found APACHE II and SOFA scores were relatively low in most enrolled patients. This is consisted with the characteristics of this disease, i.e., only in the patients with severe lung injury, but few other organs injury. COX regression confirmed that critical type COVID-19 patients showed poor prognosis and IVIG improved their survival rate. Although IVIG does not show therapeutic effect on the whole cohort, it can be beneficial to the critical type patients. In addition, COX regression also showed that lymphopenia was the risk of poor prognosis. This observation is consistent with the previous study reported that 35%-83% COVID-19 patients showed lymphocyte count decreased, and persistent lymphopenia was related with the poor outcome [26]. However, subgroup analysis based on the lymphocyte counts did not show an improved outcome related to the IVIG intervention. The explanation is uncertain yet. Future study on the role of IVIG in regulating lymphocyte number and function is needed.

There are some limitations in present study. Firstly, the cases from these eight clinical centers may still lack sufficient representativeness. Secondly, the dose and timing of IVIG administration in each center may not be exactly consistent. And thirdly, limited by the clinical workload and situation, the evaluation of immunoglobulin effect is mainly based on the clinical manifestations, rather than direct cellular and molecular assessment, including viral load and lymphocyte activation. With the progression in recognition of COVID-19, large cases randomized control studies and more developed evaluation systems are needed to confirm the efficiency of IVIG on COVID-19 treatment.

In conclusion, the present study is the first clinical research evaluating the efficiency of IVIG treatment to critical COVID-19 patients. The data demonstrate that early application of high dose IVIG can improve the prognosis of COVID-19 patients with critical type. This study provides important information on clinical application of IVIG in treatment of SARS-CoV-2 infection, including patient selection and administration timing and dosage.

## Data Availability

The data sets used and/or analyzed during the current study are available from the corresponding author on reasonable request.

## Declarations

- **Ethics approval and consent to participate**
- The study was approved by the Research Ethics Commission of General Hospital of Southern Theater Command of PLA and the requirement for informed consent was waived by the Ethics Commission.
- **Consent for publication** All authors reviewed the manuscript and approved the publication.
- **Availability of data and materials** The data sets used and/or analyzed during the current study are available from the corresponding author on reasonable request.
- **Funding**
- This work was supported by grants from the PLA Logistics Research Project of China [18CXZ030, CWH17L020, 17CXZ008], Sanming Project of Medicine in Shenzhen (SZSM20162011) and Clinical Research Project of Shenzhen municipal health commission (SZLY2017007).

## Conflict of Interest Statements

The authors declare that they have no competing interests

## Acknowledgements

We thank Prof. Jie Fan (Center for Biosecurity, University of Pittsburgh, USA) and Prof. Tianqing Peng (Center of Critical Illness Research, Western University, CAN) for revising the manuscript.

## Contributors

All authors had full access to all the data in the study and take responsibility for the integrity of the data and the accuracy of the data analysis. LZ, WM, SL, FY and HZ were responsible for study concept and design. SZ, WM, XQ, LH, GZ, LZ, LZY ZL and WC were responsible for collecting the data. WM, LZ, LZY ZL and WC were responsible for statistical analysis. LZ, WM, JJ and ZJ were responsible for drafting the manuscript.

## Supplementary

### Table legends

Table 1: Clinical parameters after the treatment with IVIG and Non-IVIG

Table 2: Effects of IVIG on clinical parameters in subgroups of critical and severe type

Table 3: Primary outcome analysis in different subgroups with IVIG treatment or not

Table 4: Effects of IVIG treatment on secondary outcome and clinical parameters in different subgroups

Table 5: Effects of different doses of IVIG treatment on clinical parameters in all patients

Table 6: Effects of different doses of IVIG on clinical parameters in different patients

Table 7: Effects of the timing with IVIG treatment on clinical parameters in all patients

### Figure legends

Figure 1: Effects of IVIG treatment on 60-day mortality in all patients

Figure 2: Effects of IVIG treatment on 60-day mortality in patients with critical type

Figure 3: Effects of IVIG treatment on 60-day mortality in patients with severe type

Figure 4: Effects of dose of IVIG treatment on 60-day mortality in all patients

Figure 5: Effects of dose of IVIG treatment on 60-day mortality in patients with critical type

Figure 6: Effects of dose of IVIG treatment on 60-day mortality in patients with severe type

Figure 7: Effects of timing of IVIG treatment on 60-day mortality in all patients

## Notes

### Competing Interest Statement

The authors have declared no competing interest.

### Clinical Trial

NA

